# Abnormalities in core AD biomarkers precede inflammatory and glial markers in CSF in Autosomal Dominant Alzheimer’s Disease

**DOI:** 10.64898/2026.03.31.26349851

**Authors:** Wenjing Lin, Aleksandra Beric, Julie K. Wisch, Bryce Baker, Gina Jerome, Matthew Minton, Sam Preminger, Jennifer Stauber, Suzanne E. Schindler, Jeffrey L. Dage, Ricardo Allegri, David Aguillon, Tammie Benzinger, Jasmeer Chhatwal, Alisha Daniels, Gregory S. Day, Emma Devenney, Nick C. Fox, Alison Goate, Brian A. Gordon, Nicolas R. Barthélemy, Jason Hassenstab, Edward Huey, Takeshi Ikeuchi, Suman Jayadev, Mathias Jucker, Takanobu Ishiguro, Jae-Hong Lee, Allan I. Levey, Johannes Levin, John C Morris, Richard J. Perrin, Alan Renton, Jee Hoon Roh, Chengjie Xiong, Randall J. Bateman, Beau M. Ances, Carlos Cruchaga, Celeste M. Karch, Charlene Supnet-Bell, Jorge Llibre-Guerra, Eric McDade, Dominantly Inherited Alzheimer Network, Laura Ibanez

## Abstract

**BACKGROUND:** Increasing evidence suggests that accurate prediction of Alzheimer’s disease (AD) symptom onset requires more than amyloid- and tau-centric biomarkers such as cerebrospinal fluid (CSF) Aβ42/40, total tau and p-tau181 and plasma p-tau217. Autosomal dominant AD (ADAD), caused by pathogenic *PSEN1, PSEN2* and *APP* mutations with predictable age at symptom onset, presents a unique opportunity to characterize the chronological changes in proteins beyond amyloid and tau and clarify them as early biomarkers of disease onset or as biomarkers related to disease staging and progression monitoring.

**METHODS:** We measured 972 CSF samples corresponding to 484 participants of the Dominantly Inherited Alzheimer Disease Network (DIAN) using the NULISASeq 120 CNS Disease Panel. We first benchmarked the technology against gold-standard measurements followed by the identification of proteins that were differentially abundant in relation to mutation status and symptomatology. Next, we determined the chronological emergence of protein changes in relation to the estimated years to onset (EYO). Finally, we assessed whether specific protein measures improved the prediction of EYO in the ADAD.

**FINDINGS:** NULISA measurements were comparable to those previously published. We demonstrated that known early alterations in CSF amyloid and tau were followed by inflammatory and neurodegenerative responses suggesting that clinical manifestation of AD happens before the inflammatory processes is fully developed. Finally, we found a multi-protein composite approach for predicting EYO that outperformed single biomarker values.

**INTERPRETATION:** Our results suggest that the main CSF proteomic landscape changes in ADAD are due to the presence of a pathogenic mutation and occur prior to symptom onset. Improved performance of multi-protein composite to predict EYO compared to single biomarker values highlights the added value of multiplex proteomic signatures for biomarker panel development.

**FUNDING:** National Institute on Aging, Alzheimer’s Association, German Center for Neurodegenerative Diseases, Raul Carrea Institute for Neurological Research, Japan Agency for Medical Research and Development, Ministry of Health & Welfare and Ministry of Science and ICT, Republic of Korea, Spanish Institute of Health Carlos III.

## INTRODUCTION

Alzheimer’s disease (AD) is the leading cause of dementia.^1^ Global dementia prevalence is estimated to be 57.4 million^2^; combined with an Alzheimer’s proportion of 60–80%^1^ suggests that over 40 million people are currently living with AD neuropathologic changes worldwide. AD pathology is characterized by the accumulation of extracellular plaques containing amyloid β (Aβ) peptide and “intracellular neurofibrillary tangles composed of phosphorylated tau protein.^3^ Autosomal dominant AD (ADAD), which accounts for less than 1% of all AD cases, has substantially contributed to our understanding of AD pathobiology. ADAD is caused by mutations in the *Amyloid Precursor Protein (APP), Presenilin-1 (PSEN1)*, or *Presenilin-2 (PSEN2)* genes, which typically result in the early onset of symptoms (generally around the age of 30-50 years).^4^ Owing to the nearly full penetrance of ADAD mutations, the age of onset in mutation carriers can be predicted based on family history and mutation,^5,6^ and this predictability makes ADAD an ideal model for studying early disease progression in presymptomatic phases.

Despite the availability of FDA-cleared biomarkers of cerebral amyloidosis, early pathologic diagnosis of AD, especially before symptom onset, remains a critical challenge for effective intervention and prevention of dementia. Early CSF biomarker studies reported that amyloid beta 42 (Aβ42)^7^, total tau (t-tau), and phosphorylated tau at serine 181 (p-tau181), show altered levels before the estimated age at symptom onset in ADAD participants.^8^ Neuroinflammation and synaptic loss co-occur with AD pathology. Studies in sporadic AD (sAD) and ADAD have reported elevation in synaptic biomarkers SNAP25 and NRGN, as well as inflammatory biomarkers GFAP and sTREM2, prior to symptom onset.^9–11^ Single analyte studies in ADAD report SNAP25 changes very early in the disease course (15–20 years prior to symptom onset), while NRGN is moderately elevated across the disease span.^10,12,13^ GFAP elevates between 10 and 15 years prior to symptom onset^11^, while changes in CSF sTREM2 levels occur after amyloid deposition and alterations in other brain injury markers, not elevating until about 5 years prior to symptom onset.^9^ Proteomic studies support these findings, demonstrating that synaptic changes occur 15 – 20 years prior to symptom onset, followed by changes in markers associated with axonal integrity and immune activation. Markers of synaptic and neuronal loss elevate near the onset of symptoms.^14–16^ In fact, the ratio YWHAG/NPTX2 in CSF is a strong predictor of cognitive decline in ADAD and sAD.^17^

Although CSF or plasma biomarker levels have been leveraged to assess brain pathology at the time of fluid sample collection, amyloid positron emission tomography (PET) remains the reference standard for evaluation of AD pathology. However, fluid biomarkers present a more cost effective and more convenient means to diagnose and monitor AD.^18^ Recent work has highlighted plasma p-tau217 as the most promising biomarker for concordance with amyloid PET.^19–23^ Thus, understanding these biomarker trajectories and their rate of change may provide crucial insights for early diagnosis and therapeutic development and intervention. Ongoing research is focused on expanding the understanding of biomarkers beyond Aβ to identify biomarker patterns associated with different stages of the disease and evaluating their potential use in clinical practice, as the lack of understanding of the interaction between biomarker trajectories, genetics and symptom onset, makes it difficult to interpret clinical trial results or predict individual responses to treatment, especially in the context of amyloid removal therapy.

Nucleic acid linked immuno-sandwich assay (NULISA^™^) is a mid-throughput method, with 125 proteins available in the NULISAseq^™^ CNS Disease Panel, that requires low sample volumes and has multiplexing capabilities.^24^ An increasing number of studies utilizing NULISAseq^™^ CNS Disease Panel have shown it to provide dependable measurements for established AD biomarkers in both plasma and CSF,^25,26^ that highly correlate with immunoassay and IP-MS^25^, and associate with amyloid-PET status.^25–27^ However, most of these studies focus primarily on sAD and core AD biomarkers, thus missing out on the full potential of the NULISAseq^™^ CNS Disease Panel, which we are leveraging to assess all the proteins on this panel at much earlier stages than has been possible with sAD to provide novel insight to earlier pathogenic changes that go beyond the core biomarkers and would not be discoverable otherwise.

In this study, we leveraged longitudinal data from the Dominantly Inherited Alzheimer Network (DIAN), natural history study of ADAD caused by *APP, PSEN1*, and *PSEN2* mutations, to study the chronology of AD biomarkers in CSF using the NULISA technology. We first benchmarked the use of this technology in an ADAD cohort by testing the correlation between immunoassay or IP-MS and NULISA measurements for core AD biomarkers (Aβ42/40 ratio, Aβ42, Aβ40, p-tau181, p-tau217, and total tau), other markers of neurodegeneration/neuronal damage (NFL, NRGN, VILIP1, and SNAP25), as well as inflammatory (GFAP and YKL40) proteins in CSF that were already available in the DIAN study. Then, we identified differentially accumulated proteins and explored the timing of abnormal changes in the proteins included in the NULISAseq CNS Disease Panel v1. Finally, we built multi-protein predictive models to improve the estimation of time to onset.

## METHODS

### DIAN Summary Assessment

DIAN is an international observational study that focuses on the recruitment of families with AD that show an autosomal dominant pattern of inheritance caused by mutations in *PSEN1, PSEN2, or APP*. The study uses standardized assessments to collect longitudinal data, including clinical, cognitive, imaging, and fluid biomarker measurements.^28^ Participants were recruited through the DIAN Observational Study (ClinicalTrials.gov Identifier: NCT00869817) and have provided written consent or assent with proxy consent prior to enrollment in accordance with the latest Helsinki Declaration. The study was approved by the Human Research Protection Office and the Institutional Review Board (IRB) at Washington University in St Louis, USA and the respective participating sites. Study participants were enrolled from 2008 to 2023.

DIAN participants are assessed comprehensively every two years while asymptomatic (CDR=0), and annually at −5 years before expected symptom onset or once they become symptomatic (CDR>0). Assessments are performed by clinicians who are blinded to participants’ genetic status and include: reviews of family and personal medical history, clinical interview, a detailed neurological examination and cognitive assessments, the Geriatric Depression Scale (GDS), and the Neuropsychiatric Inventory (NPI). Dementia severity is determined using CDR, following standardized protocols. Mutation status used to be determined using a PCR-based amplification of the relevant exon, followed by Sanger sequencing, or via short-read whole genome and/or whole exome sequencing.^7,28^ The Estimated Years to Onset (EYO) is one of the key variables for DIAN participants. It is recalculated every year based on the age at last visit and the estimated dementia symptom onset age specific to their mutation.^5–7,29^

### Study participants

*We* included 972 CSF samples that had NULISAseq™ CNS Disease Panel v1 data available, from 484 unique DIAN participants (Table 1). All participants were of European ancestry, and 56% of the participants were female. Mean age at first visit was 37.25±10.72 years, with a mean follow up time of 3.73±2.42 years, 66% were mutation carriers, and 36% of those were already symptomatic at the time of CSF collection. There was no significant difference in age at draw between Mutation Carriers (MC) and non-carriers (NC; p=0.12).

**Table 1.** Baseline demographic characteristics. Baseline draw was used for those participants with more than one time point available. P value was calculated for the comparison of MCs to NCs using t-tests for continuous variables and chi-square tests for categorical variables.

| Sample Type | Total (n=484) | Mutation Carriers (n=321) | Non-Mutation Carriers (n=163) | P value |
| --- | --- | --- | --- | --- |
| Sex (n, % females) | 275 (57%) | 181 (56%) | 94 (58%) | 0.94 |
| Age at draw (years, mean±sd) | 37.26±10.72 | 37.80±10.55 | 36.16±11.09 | 0.12 |
| EYO (years, mean±sd) | - | -8.72± 10.91 | -11.59± 11.49 | - |
| Follow-up time (years, mean±sd) | 3.73±2.42 | 3.62±2.54 | 4.03±2.11 | 0.18 |
| Symptomatic at draw (n, %) | 106 (22%) | 97 (30%) | 0 (0%) | - |
| Mutation carriers (n, %) | 321 (66%) | 321 (100%) | 0 (0%) | - |
| PSEN1 carriers (n, %) | - | 248 (77%) | - | - |
| PSEN2 carriers (n, %) | - | 21 (6%) | - | - |
| APP carriers (n, %) | - | 52 (16%) | - | - |

n=sample size; sd=standard deviation; EYO=Estimated Years to Onset; PSEN1=Presenilin 1; PSEN2=Presenilin 2; APP=Amyloid Precursor Protein

### Protein measurements

CSF immunoassay Aβ40, Aβ42, t-tau, and p-tau181 measurements were obtained using a Lumipulse G1200 Chemiluminescent Enzyme Immunoassay Assay platform as previously described.^18,30,31^ CSF NRGN, SNAP25, and VILIP1 levels were measured using Single Molecule Counting (SMC) technology (EMD Millipore, Burlington, MA)^32–35^ while Chitinase 3 Like 1 (YKL40) CSF concentration was measured using the MicroVue YKL40 EIA plate-based enzyme immunoassay (Quidel, San Diego, CA) for serum and plasma kit.^36^ Additionally, Aβ40, Aβ42, p-tau181, and p-tau217 concentrations were measured using immunoprecipitation-mass spectrometry (IP-MS) as previously described.^37–40^ Finally, CSF soluble TREM2 (sTREM) was quantified using an in-house Meso Scale Discovery (MSD)-based immunoassay.^41^

Samples were also quantified using the NULISAseq™ CNS Disease Panel v1, following a previously described protocol.^25^ In short, target proteins in the sample were incubated with DNA conjugated antibodies to form immunocomplexes. The resulting immunocomplexes were purified via a series of steps with oligo-dT and then with streptavidin beads, followed by proximity ligation to produce DNA reporters. DNA reporters were subsequently PCR amplified, yielding next-generation sequencing libraries, which were sequenced by PrimBio Research Institute, LLC. Detected protein levels are normalized using internal and inter-plate controls, then log_2_ transformed, supplying the reported NULISA Protein Quantification (NPQ) values.

### Quality Control

Prior to analysis, immunoassay (ELISA and SMC) and IP-MS obtained protein concentrations were log_2_ transformed to approximate to a normal distribution. The NULISAseq™ CNS Disease Panel v1 reports values as NPQ units. These units are already standardized, normalized and log_2_ transformed. Outliers were identified using the Interquartile Range (IQR) method and removed if they were outside the *Q1-1.5×IQR-Q3+1*.*5×IQR* range from the mean. Samples that were below the detectability threshold, defined as a minimum of 70% of target proteins being above the level of detection, were removed. NULISA quantification was produced across multiple assay plates, plate-level technical variation was observed between runs. To account for this, measurements were scaled within each plate prior to downstream analyses to reduce plate-to-plate differences. Finally, we performed principal component analyses (PCA) and further removed any outliers defined as any samples that were more than three standard deviations removed from the means of PC1 or PC2 (Supplementary Figure 1A).

### Benchmarking of NULISA in CSF from DIAN participants

Spearman correlation analyses were carried out between all NULISA and immunoassay/IP-MS/SMC measurements available. P-values were adjusted for multiple testing using the Benjamini–Hochberg false discovery rate (FDR). Corrected p-values below 0.05 were considered significant.

### Identification of altered proteins

To identify proteins that were differentially abundant between groups, we leveraged a cross-sectional sub-sampling that only included the most recent visit for each participant (Supplementary Table 1). We performed several pair-wise comparisons: (i) mutation carriers (MC) vs non-mutation carriers (NC); (ii) a comparison of each ADAD gene, *PSEN1, PSEN2* or *APP*, carriers to non-carriers, and (iii) symptomatic mutation carriers (sMC) vs asymptomatic mutation carriers (aMC). All differential abundance analyses adopted a cross-sectional approach, utilizing only one observation per participant and *lm* function from the *stats* R package for logistic regression, and were adjusted by EYO and sex. All p-values were corrected for multiple testing using Benjamini-Hochberg method; corrected p-values lower than 0.05 considered significant.

### Chronological changes of protein abundance

To assess the chronological order in which proteins change in relation to EYO, we modeled the probability of protein profile abnormality as a function of EYO in MC. Abnormality thresholds were defined using NC distributions by examining the protein distributions in NCs and MCs.^7^ We selected the 90th percentile for protein levels considered to increase with disease progression (Supplementary Figure 2A) and the 10th percentile for those considered to decrease with progression (Supplementary Figure 2B), since these correspond to the percentiles that best separated the two groups. Each biomarker measurement in each sample was dichotomized as abnormal or normal based on these thresholds. We fitted logistic mixed-effects models using the *glmer* function from the *lme4* package in R to estimate abnormality probability as a function of EYO in MCs. Models included EYO as a fixed effect and random intercepts for individual subjects to account for within-subject correlation from repeated measurements. This process was repeated for all proteins included in the NULISASeq CNS panel. Results were shown only for those proteins that had at least 50% probability of reaching abnormality in MC.

Amyloid beta measurements are affected by the number of freeze/thaw cycles.^42–44^ The samples included in this work had a diverse number of freeze/thaw cycles, which made us suspect potential protein drift. Thus, we compared the abnormality curves of Aβ40/42 and p-tau217 when measured using CSF with only one freeze/thaw cycle to inform if Aβ was affected by protein drift, using p-tau217 as control. We observed that Aβ40/42 levels measured with NULISA changed later than those measured with IP-MS or Lumipulse probably confirming the suspected protein drifting (Supplementary Figure 1B), whereas p-tau217 was not affected (Supplementary Figure 1C). Thus Aβ40/42 was removed from this analysis and replaced by IP-MS measurements to avoid protein drift effects.

### Improvement of EYO prediction with XGBoost

We constructed gradient-boosted decision tree models using the XGBoost algorithm (eXtreme Gradient Boosting) to predict EYO from CSF protein measures. All models were trained on 60% of the data, with 40% held out for testing. We excluded the subset of participants who became symptomatic in the DIAN study (n=24) and leveraged them for subsequent sensitivity analyses. To ensure the statistical independence required by XGBoost, and to get most advanced pathological state possible, only the most recent visit for each participant was considered (Supplementary Table 1).

We compared the performance of four models: (i) age, (ii) individual AD-core biomarkers, (iii) PET-PiB amyloid imaging, and (iv) a multiprotein model that relied on LASSO (L1 regularization) regression for feature selection. Then, we conducted a sensitivity analysis leveraging the 24 individuals undergoing clinical conversion to assess the models’ ability to predict proximity to real symptom onset.

To identify proteins for inclusion in the multiprotein model, we performed a 50-fold bootstrap validation. In each iteration, we randomly sampled 60% of the training data and fit a LASSO model with regularization parameter λ optimized via 10-fold cross validation at each iteration. We ranked the protein by how many times each was selected as informative by LASSO across all iterations. Proteins included in at least 12% of bootstrapped iterations were included in the final combined model. Subsequently, we evaluated XGBoost models with incrementally increasing numbers of top-ranked proteins via 5-fold cross-validation on the training set and selected the optimal set by minimizing mean absolute error (MAE). Hyperparameters of the XGBoost were optimized via grid search with internal validation. Models were trained to minimize a squared-error objective function, utilizing an early stopping protocol (patience=50 rounds) to mitigate overfitting. The predictive models were evaluated on the independent test set (40% of the cross-sectional cohort) using mean absolute error (MAE). We compared the performance of each model relative to an age-only model (null) to assess whether CSF proteins provided additional predictive value. To evaluate model performance across the full range of EYO, we applied point-wise paired bootstrap analysis at each observed EYO. We calculated the difference in MAE at each EYO observation and applied a 1,000-iteration paired bootstrap to estimate the corresponding 95% confidence intervals.

## RESULTS

### NULISA quantification of AD biomarkers in the CSF of DIAN participants correlates with established methods

We compared CSF Aβ40, Aβ42, total tau, and p-tau181 values measured using NULISA and Lumipulse or IP-MS (Fig1A, STable2). Aβ showed strong and highly significant correlations when measured NULISA and Lumipulse (Aβ40 (ρ=0.84, p=1.09×10^−264^); Aβ42 (p=0.88, ρ=1.98×10^−323^); Aβ42/40 ratio (p=0.87, p=5.48×10^−295^)), or IP-MS (Aβ40 (ρ=0.84, ρ=2.11×10^−159^); Aβ42 (ρ=0.87, ρ=2.58×10^−185^); Aβ42/40 ratio (ρ=0.88, p=1.79×10^−190^)). As expected, the correlations between Lumipulse and IP-MS were strong (Aβ40: ρ=0.92, p=1.89×10^−233^;Aβ42: ρ=0.94, p=1.64×10^−278^; Aβ42/40: ρ=0.94). We obtained very similar results for the tau species, with the strongest correlation observed for p-tau181 (ρ=0.92, p<2.2×10^−308^) followed by total tau (ρ=0.83, p=4.38×10^−222^) when comparing Lumipulse and NULISA. IP-MS p-tau181 and p-tau217 showed correlations of 0.91 (p=4.41×10^−258^) and 0.91 (p=6.78×10^−253^), respectively, between IP-MS and NULISA.

**Figure 1.**
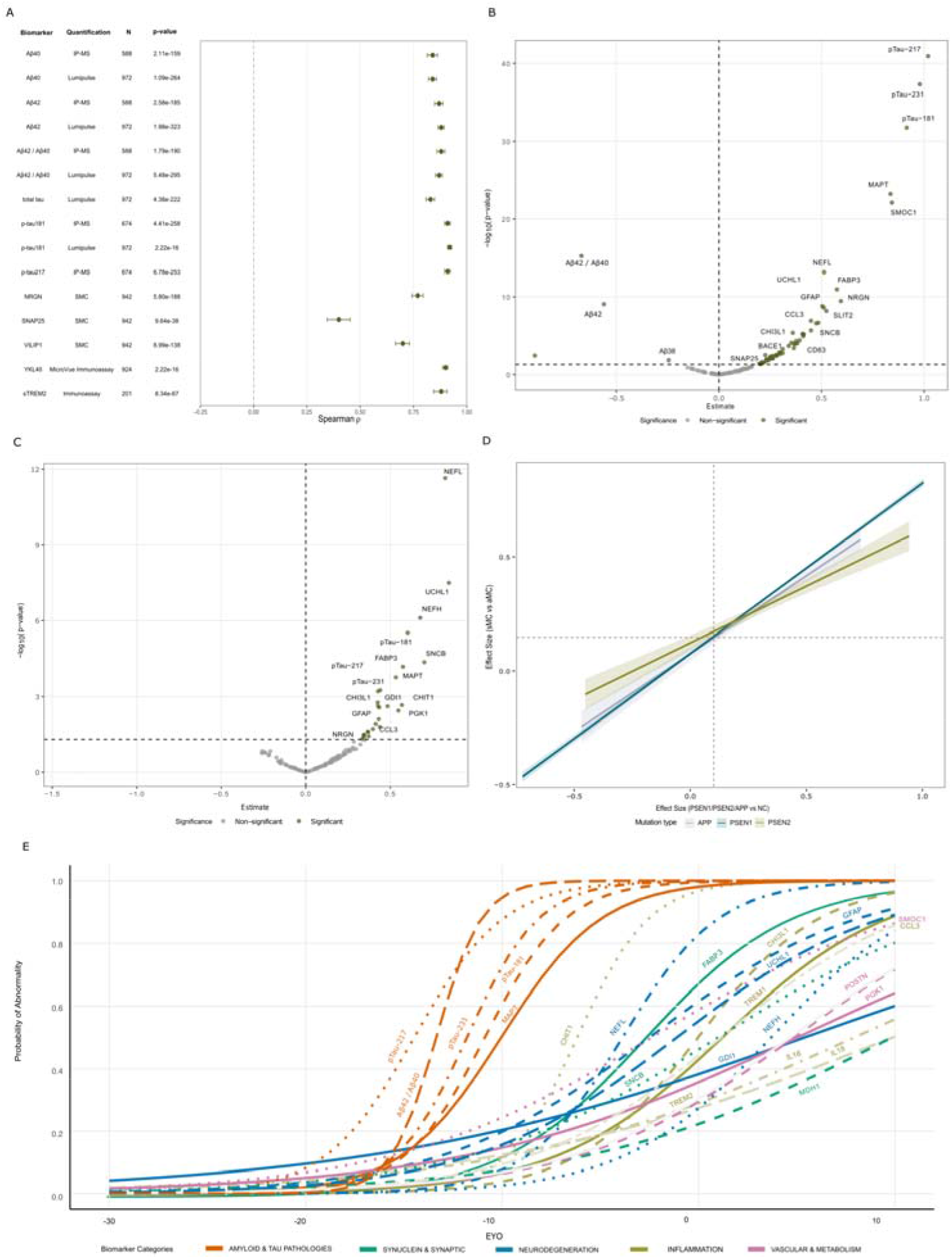
NULISA captures proteomic signatures in CSF. **A**. Benchmarking of NULISA measurements in CSF of DIAN participants (raw p-values are listed) B. Volcano plot showing the results of comparing CSF measurements in Mutation Carriers (MC) vs Non-Carriers (NC) at last visit **C**. Volcano plot showing the results of comparing CSF measurements in Symptomatic Mutation Carriers (sMC) vs asymptomatic Mutation Carriers (aMC) at last visit D. Effect size correlation for the results of the Mutation Carriers vs Non-Carriers comparison and the *PSEN1,PSEN2,APP* Carriers vs Non-Carriers comparison. E. Abnormality curves for all NULISA measurements that reach 50% abnormality; legend is sorted in order that they reach abnormality.

Additional biomarkers, YKL40, NRGN, VILIP1, and SNAP25, were measured using both traditional immunoassay and NULISA. All of them showed significant correlations between the two approaches, YKL40 (ρ=0.90, p<2.2×10^−308^), NRGN (ρ=0.77, p=5.80×10^−188^), and VILIP1 (ρ=0.70, p=8.99×10^−138^) (Supplementary Table 2, Figure 1A). SNAP25 was only moderately correlated (ρ=0.40; p=9.64×10^−38^). Finally, CSF sTREM2 was quantified by immunoassay and showed strong correlation with the NULISA measurements (ρ=0.88, p=8.34×10^−67^).Overall, we showed that NULISA measurements strongly correlate to the established immunoassay and IP-MS methods in the DIAN study as previously described for sAD.^25^

### CSF Proteome Reveals Widespread Proteomic Alterations in ADAD, regardless of symptomatology or affected gene

Out of 125 proteins successfully quantified in the last CSF visit of 484 DIAN participants (321 MCs and 163 NCs) using NULISA technology, 61 were associated with ADAD pathogenic mutation status independent of symptomatology (Figure 1B), whereas 15 were associated with clinical symptoms within MCs (Figure 1C; Supplementary Table 3). This suggests that in this population many of the changes in the proteomic landscape identify the asymptomatic stage of disease in pathogenic mutation carriers. We then stratified by affected ADAD gene to examine any potential differences due to the gene groups, (Supplementary Table 3). When *PSEN1* carriers were compared to NC, the effect sizes of the two previously tested analyses were highly correlated (ρ=-0.86, p=3.20×10^−39^). Differences in p-values were most likely driven by differences in sample size. Similar results were observed when comparing *PSEN2* (ρ=-0.09, p=0.34) or *APP* (ρ=-0.22, p=0.01) carriers to non-carriers (Figure 1D). These results suggest that mutation in these three genes drive changes that are similar regardless of the affected ADAD gene.

Dysregulated proteins with strongest associations with mutation carrier status have already been described and include the phosphorylated tau species, p-tau217 (effect size=1.02, p=1.20×10^−41^), p-tau231 (effect size=0.98, p=4.93×10^−38^), and p-tau181 (effect size=0.91, p=1.77×10^−32^), followed by total tau (MAPT; effect size=0.84, p=6.00×10^−24^), SMOC1 (effect size=0.84, p=6.00×10^−24^), and Aβ42/40 ratio (effect size=-0.67, p=5.84×10^−16^). However, when comparing aMC to sMC the top proteins were neurofilament light (NFL; effect size=0.83, p=2.26×10^−12^), ubiquitin C-terminal hydrolase 1 (UCHL1; effect size=0.85, p=3.22×10^−08^) and neurofilament heavy polypeptide (NEFH; effect size=0.68, p=7.8OxlO’^07^); all known to be associated with neuronal health,. These proteins were also significant in the MC vs NC comparison, but ranked seven, twelve, and 37 respectively. The Aβ42/40 ratio was not significant in the sMC vs aMC comparison and the phosphorylated tau species ranked five (p-tau181), eight (p-tau 231), and nine (p-tau217). Altogether, these observations support the knowledge that amyloid biomarker abnormalities are present before the onset of symptoms.

The NULISAseq^™^ CNS Disease Panel v1 measures 52 inflammatory proteins, some of which have been previously associated with AD or ADAD. Among them, CCL3^45^ was significant in both comparisons, MC vs NC and sMC vs aMC (effect size=0.45, p=1.19×10^−7^; effect size=0.43, p=2.69×10^−3^), while sTREM2^9^ was only significant in the MC vs NC comparison (effect size=0.31, p=5.43×10^−4^) along with TREM1^16^ (effect size=0.39, p=3.88×10^−5^), IL15^16^ (effect size=0.26, p=4.99×10^−3^), and SLIT2^16^ (effect size=0.52, p=6.43×10^−9^) suggesting that not all inflammatory alterations caused by the presence of a mutation precede symptom onset. There were other proteins, not previously reported in the context of AD or ADAD, such as, TIMP3 (effect size=0.38, p=4.92×10^−5^) and IL16 (effect size=0.34, p=2.10×10^−4^).

Among the other proteins included in the NULISA panel, FABP3 (effect size=0.57, p=l. 11×10^−11^), PARK7 (effect size=0.25, p=7.94×10^−3^), SOD1 (effect size=0.37, p=1.13×10^−4^), VSNL1 (effect size=0.47, p=2.51×10^−7^), and UCHL1 (effect size=0.51, p=2.36×10’^9^) were significantly higher in MC compared to NC. Several synuclein species were also found to be associated with mutation status: α-synuclein (effect size=0.31, p=1.77×10^−3^), α-synuclein phosphorylated at serine 129 (pS129-α-syn^38^; effect size=0.41, p=1.14×10^−5^) and β-synuclein (effect size=0.48, p=2.18×10^−7^), which was also associated with the presence of symptoms (effect size=0.70, p=4.37×10^−5^).

### Markers of inflammation change approximately six years before the onset of symptoms with the dysregulation of Chitotriosidase 1

*\Ne* calculated protein changes as a function of EYO in MC to chronologically describe when the level of each pathological protein reaches a 50% probability of being abnormal relative to NCs (Figure 1E). As previously described^46,47^, we observed that core AD biomarkers (p-tau217, Aβ42/40, p-tau181, and total tau) reach a 50% probability of being abnormal first, between 20 and 10 years before symptom onset (Supplementary Table 4). CHIT1 reaches this threshold about six years before onset followed by NFL (four years before onset), FABP3, GFAP, and SMOC1 (about two and a half years before symptom onset). YKL40 reaches 50% probability to be abnormal right at the onset of symptoms. After symptom onset, several inflammatory proteins reach this same threshold in the span of approximately 10 years including TREM1, CCL3, TREM2, IL6 or IL18, suggesting that neurodegeneration of AD (manifesting as symptoms) becomes significant even before the inflammatory processes have fully developed.

The quantification of proteins reaching the 50% probability of being abnormal within the EYO available in the DIAN study were highly correlated, suggesting parallel trajectories (Supplementary Figure 4A and Supplementary Table 5). Total tau and phosphorylated tau forms showed the highest correlations, with ρ>0.90. Other pairs showed correlations of more than 80% such as FABP3 and total tau (MAPT in the NULISA panel); FABP3 and MDH1, NFL and YKL40 (CHI3L1), SNCB and MDH1, or SNCB and FABP3, suggesting coordinated mechanisms for protein abundance regulation such as funnel effects or master regulators. The Aβ42/40 was weakly correlated with other protein measurements. It showed the highest correlation with p-tau217 (ρ=0.65) and other tau phosphorylated species and NFL (ρ=-0.59). Given the high correlation observed, we performed gene-enrichment analyses (STab6 and SFig4B) and found one KEGG pathway enriched *(neurodegenerative disease*, p=4.55×10^−4^), and 421 GO terms that passed multiple test correction. Several GO terms identified are relevant to ADAD, *neuroinflammatory response* (p=2.77×10^−8^), *microglial cell activation* (p=2.08×10^−7^), *positive regulation of inflammatory response* (p=6.42×10^−7^), and *positive regulation of neuroinflammatory response* (p=8.31×10^−7^) among many others.

### Multiprotein models improve the prediction of EYO compared to established biomarkers

*\Ne* applied machine learning to CSF proteomic data to identify the minimal set of proteins necessary to predict EYO in presymptomatic MCs. The best-performing model included 35 proteins. Feature importance analysis revealed that GFAP contributed most strongly to predictions (gain=0.16), followed by p-Tau217 (gain=0.08), NGF (gain=0.07), Aβ42/40 ratio (gain=0.06), and Aβ38 (gain=0.05; Figure 2A). Repeated LASSO regularization across resampled training subsets reduced redundancy among correlated predictors by selecting stable features. Pairwise correlation analysis of the selected proteins showed moderate correlations (Supplementary Table 6). The multi-protein model achieved minimal MAE at 3.96 years (Fig 2B; Root Mean Squared Error - RMSE=5.22), significantly outperforming the age-only model (Figure 2C; MAE=5.75, RMSE=7.10). When testing against amyloid imaging, the multi-protein model also outperformed amyloid imaging (Figure 2B, MAE=5.19, RMSE=6.29). We then compared the multi-protein model to the performance of already established AD biomarkers measured using IP-MS p-tau217 (Figure 2E; MAE=4.75, RMSE=5.78), p-tau181 (Figure 2B; MAE=5.66, RMSE=7.00), Aβ42/40 ratio (Figure 2B; MAE=5.30, RMSE=6.82) and the YWHAG/NPTX2 ratio (Figure 2B; MAE= 6.68, RMSE= 8.00). Point-wise paired bootstrap analysis at each EYO observation revealed that the multi-protein model significantly outperformed the null model consistently in the –17 to −4 years before onset (EYO) window (Figure 2C). Significance was determined by 95% bootstrap confidence intervals of the MAE difference excluding zero, with bootstrap p-values < 0.05 reported as supportive evidence (Supplementary Table 7). Although single biomarker models showed modest improvements compared to the age-only baseline in overall MAE, their predictive benefit was temporally limited (Figure 2C).

**Figure 2.**
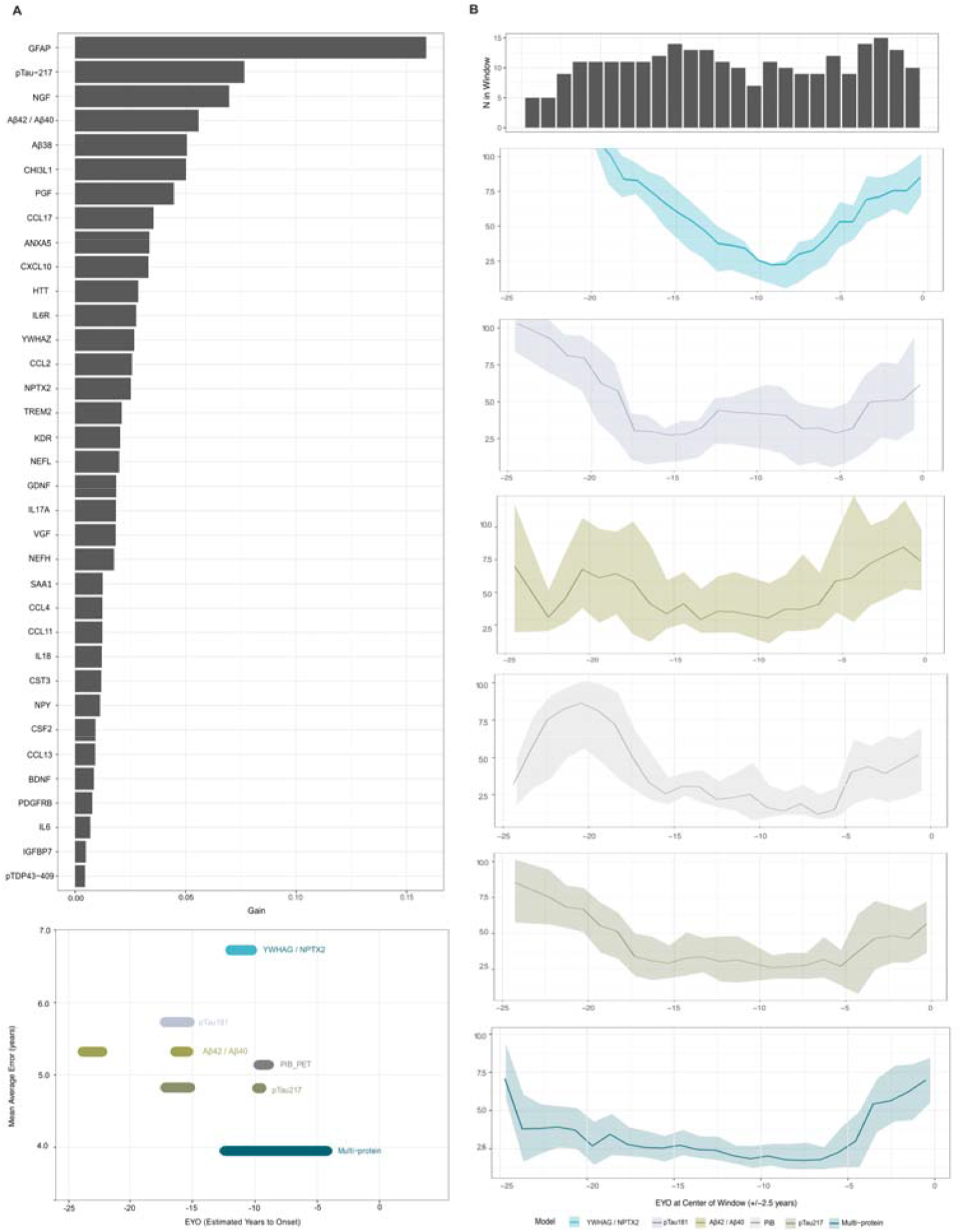
Multiprotein Model Design and EYP Prediction Performance. **A**. Feature importance in the multi-protein model ranked by gain score. B. Sample distribution and performance of the YWHAG / NPTX2, p-tau181, Aβ42/Aβ420, PET-PiB, p-tau217 and multiprotein models across EYO windows (±2.5 years). Mean absolute error (MAE) shown with 95% confidence intervals. C. EYO ranges in which each model outperformed the age-only null model. Points indicate the overall MAE of each model, and horizontal position reflects the temporal windows where the improvement was statistically significant.

To further test the multiprotein model, we assessed performance predicting actual age at onset using clinical converters (n=24) who were held out from model training (Supplementary Figure 5). The multiprotein model achieved an MAE of 7.88 (RMSE=9.11), comparable to single-biomarker models including p-tau217 (MAE=5.86, RMSE = 6.68), p-tau181 (MAE=6.86, RMSE=7.75), Aβ42/40 (MAE=7.88, RMSE=8.45), and PiB PET (MAE=6.81, RMSE=7.45). The model containing only age, also exhibited inferior performance (MAE=7.57, RMSE=9.15). Although the multi-protein model showed numerically higher MAE than single protein models in converters, bootstrap analysis showed wide confidence intervals with no statistically significant differentiation between models (Supplementary Figure 5), confirming the limited statistical power in this sub-cohort.

## DISCUSSION

In this study, we leverage the rare and unique ADAD population and NULISA measurements to describe the presymptomatic phases of AD and predict EYO. ADAD and sAD show shared pathophysiology and in consequence, our findings are most likely true in sAD. We have shown strong correlation for NULISA quantifications compared to traditional biomarker measurements and replicated previous findings in sAD.^25^ Most of the known AD biomarkers are already altered in MCs long before the onset of symptoms, suggesting that the main proteomic landscape changes are due to the presence of a pathogenic mutation and precede symptom onset. Though in this study the changes are attributed to the presence of a pathogenic mutation, they are most likely part to the pathogenic events that will ultimately cause the onset of AD, and that it is shared between ADAD and sAD. Overall, proteomic changes appeared similar across individuals who carry ADAD mutations, regardless of which gene is affected. Early alterations in tau and amyloid were followed by inflammatory and neurodegenerative responses. Finally, we found that a multi-protein composite approach to predicting EYO outperformed single biomarker values or age, highlighting the value of multiplex proteomic signatures for biomarker panel development.

In ADAD, proteomic studies show widespread CSF protein dysregulation that begins decades before symptoms, involving amyloid⍰related extracellular matrix proteins, followed by synaptic, metabolic, axonal, inflammatory and neurosecretory pathways. We were able to replicate several of those findings. For example, we added further evidence that the amyloid cascade is already dysregulated fifteen years prior to the onset of symptoms including the amyloid beta ratio^7,14,40,48^ and phosphorylated tau species.^39,40,49^ We were also able to replicate that SMOC1^14^, NFL^50−52^, VSNL1 (VILIP1)^10^ and YKL40^10^ are associated with being carrier of a mutation in *APP, PSEN1*, or *PSEN2. \Ne* identified CHIT1, elevation of which has been reported in sAD^53^ but not in ADAD. CHIT1 is a microglial enzyme that seems to promote antiinflammatory polarization, enhance amyloid beta phagocytosis, supports lysosomal function, and protects neurons in animal models.^54,55^ What we have demonstrated here is that those changes seem to be related to the presence of a pathogenic mutation that ultimately is causing dysregulation in the Ab pathway, regardless of the presence of symptomatology. However, NFL^56^ or YKL40^57–59^ are associated with the symptomatology, which aligns with the findings in sAD. Once symptomatic, other changes become more evident such as the dysregulation of NFL, UCHL1, or NEFH, suggesting that in the ADAD population those might be informative of symptom onset, as supported by the multiprotein models.

Some reported findings, however, have not been replicated in the current study. We were not able to replicate previous findings such as the associations between the levels of SNAP25^10^, NPTX1^60^, NPTX2^60^, NPTXR, or YWHAZ and mutation status.^14^ Differences in sample selection or epitope captured by each platform (NULISA, SomaLogic, or MS) might explain the difference in these findings. We were unable to test the association of YWHAG^17^, also previously reported, due to failure during quality control.

Using a novel platform, we have characterized the inflammatory landscape of ADAD. Neuroinflammatory proteins such as TREM1 and GFAP, general inflammatory proteins such as IL16 and TAFA5, as well as neurodegeneration marker such as NFL, are elevated in MCs. Temporal analyses using a 50% probability of abnormality approach showed that the changes in the levels of inflammatory proteins GFAP, YKL40, TREM1 followed changes in core AD biomarkers. This provides valuable insight into the possible order of the pathological cascade that precedes symptom onset, simultaneously providing support for further investigation of these proteins as potential early-stage biomarkers for disease detection, monitoring, and clinical benefits of targeting inflammation in symptomatic and presymptomatic individuals.

Reflecting the central role of APP processing in AD pathology, we found that CD63 and BACE1 levels were elevated relative to mutation, but not symptom status. BACE1 is a β-secretase responsible for the production of a soluble APPβ fragment, which is further processed by γ-secretase, giving rise to Aβ peptides.^41^ CD63 might aid in directing APP and BACE1 to late endosomes and lysosomes, thus directing APP toward amyloidogenic cleavage. Our observations support previous reports on the importance of BACE1 for APP processing and provide evidence that this processing is helped by CD63, despite some reports suggesting that APP and CD63 are sorted into different exosome populations.^42^

Overall, when stratifying by ADAD gene, we did not find striking differences. The differences in p values were most likely driven by differences in sample size given the correlations observed in effect sizes. Similar results were observed when comparing *PSEN2* or *APP* carriers to non-carriers. These results suggest that mutation in these three genes drive changes that are similar regardless of the affected ADAD gene. That does not mean that there are no differences in the proteomic profile of carriers of different mutations, but suggests the differences are more subtle and studies with larger sample sizes are required to have enough statistical power to identify them with certainty.

Finally, we leveraged CSF data to develop machine learning models capable of predicting time-to-onset in mutation carriers. The purpose of this analysis was primarily inferential rather than predictive. Previous work demonstrated that amyloid positivity timing explains 39% of EYO variance in ADAD^61^, suggesting that inflammatory markers in our proteomic panel change more proximally to symptom onset and provide more informative temporal predictions. When we consider feature importance, we find that GFAP, p-tau217, and NGF are most vital to predicting the temporal progression of the disease, with GFAP contributing most to the model, which is likely explained by the fact that GFAP exhibits the most gradual and steepest change leading up to the onset of symptoms. This suggests that changes in neuroinflammation, in addition to traditional amyloid pathology, are associated with preclinical disease progression, consistent with prior work using other proteomic platforms^16^ or focusing on LOAD. This finding is interesting in light of earlier work that pooled multiple single analyte assays to predict EYO and found that CSF biomarkers of neuronal integrity and inflammation (NFL, NRGN, SNAP25, VILIP1 and YKL40) offered relatively little information in the prediction of EYO.^10,12^ This may suggest that GFAP and NGF are more specifically associated with AD progression than these other biomarkers of neuronal integrity and inflammation. Other common biomarkers that are used to describe AD progression like p-tau181 and p-tau217 were less informative than the multiprotein panel. Notably, both of these outperformed the null model around EYO=-15, suggesting that tau phosphorylation changes occur in this time range, as previously proposed.^8,40^ Altogether, machine learning models integrating multiple CSF biomarkers robustly predicted time-to-symptom onset outperforming traditional single-marker approaches and amyloid imaging, highlighting the potential of multiprotein signatures for forecasting disease progression in ADAD.

Despite being the latest and largest longitudinal assessment of the CSF proteome in DIAN, this study has several limitations. Despite the limited sample size, this is the largest ADAD population to date. The NULISA panel is a curated panel to include known proteins, limiting our findings and conclusions to the proteins quantified. The NULISA CNS panel does not provide absolute quantifications, limiting the current clinical interpretability and the identification of thresholds for potential multiprotein EYO prediction, despite providing evidence on the use of multiprotein biomarkers.

In conclusion, this study provides a comprehensive quantitative characterization of CSF proteomic changes across the ADAD continuum using the NULISA platform, validating its robustness and translational value for biomarker discovery. By integrating cross-sectional analyses with genetic stratification and machine learning, we delineate a temporal cascade of protein level changes—from early amyloid and tau changes to subsequent inflammatory and neurodegenerative processes—and reveal shared dynamics across *APP, PSEN1*, and *PSEN2* mutation carriers. The predictive performance of multi-protein models highlights the promise of multiplex proteomic signatures for guiding precision medicine approaches in ADAD, with potential applicability to sAD.

## DATA AVAILABILITY

Data and study samples used in this study are available upon request and will follow the policies of the DIAN (https://dian.wustl.edu), which comply with the guidelines established by the Collaboration for Alzheimer’s Prevention.

## ACKNOWLEDGMENTS

We acknowledge the altruism of the participants and their families and contributions of the DIAN research and support staff at each of the participating sites for their contributions to this study, without whom this study would not have been possible. Data collection and sharing for this project was supported by The Dominantly Inherited Alzheimer Network (DIAN, U19AG032438) funded by the National Institute on Aging (NIA), the Alzheimer’s Association (SG-20-690363-DIAN), the German Center for Neurodegenerative Diseases (DZNE), Raul Carrea Institute for Neurological Research (FLENI), Partial support by the Research and Development Grants for Dementia from Japan Agency for Medical Research and Development, AMED, the Korea Health Technology R&D Project through the Korea Health Industry Development Institute (KHIDI), Spanish Institute of Health Carlos III (ISCIII), Canadian Institutes of Health Research (CIHR), Canadian Consortium of Neurodegeneration and Aging, Brain Canada Foundation, and Fonds de Recherche du Québec Santé. Additionally, the analysis were supported by grants from the National Institutes of Health AG062723 and P01AG003991, the Michael J Fox Foundation, the Alzheimer’s Drug Discovery Foundation, Bright Focus Foundation, and the Department of Defense and made possible thanks equipment made possible by the Hope Center for Neurological Disorders, the Neurogenomics and Informatics Center (NGI: https://neurogenomics.wustl.edu/) and the Departments of Neurology and Psychiatry at Washington University School of Medicine.

This manuscript has been reviewed by DIAN Study investigators for scientific content and consistency of data interpretation with previous DIAN Study publications.

## FUNDING

Data collection and sharing for this project was supported by The Dominantly Inherited Alzheimer Network (DIAN, U19AG032438) funded by the National Institute on Aging (NIA), the Alzheimer’s Association (SG-20-690363-DIAN), the German Center for Neurodegenerative Diseases (DZNE), Raul Carrea Institute for Neurological Research (FLENI), Partial support by the Research and Development Grants for Dementia from Japan Agency for Medical Research and Development (AMED: JP25dk0207066), the Korea Health Technology R&D Project through the Korea Health Industry Development Institute (KHIDI), Korea Dementia Research Center (KDRC), funded by the Ministry of Health & Welfare and Ministry of Science and ICT, Republic of Korea RS-2024-00344521, and Spanish Institute of Health Carlos III (ISCIII). This manuscript has been reviewed by DIAN Study investigators for scientific content and consistency of data interpretation with previous DIAN Study publications. We acknowledge the altruism of the participants and their families and contributions of the DIAN research and support staff at each of the participating sites for their contributions to this study.

This work was supported by grants from the National Institute of Health (P30AG066444, R00AG062723, U19AG03243812, R01AG053267, RF1NS075321) and Alzheimer’s Association (DIAN-TU-PP-22-872356 and DIANTUOLE21725093).

## AUTHOR CONTRIBUTIONS

LI, and AB conceived this article. WL, AB, JW, and LI wrote the drafted the manuscript. LI conceptualized and designed the research plan. LI AB, and JW designed the analysis plan. AB and WL processed all the data and performed the analyses. BB, GJ, MM, JS, SP obtained and processed the samples. BB generated the NULISAseq™ CNS Disease Panel 120 v1 data. All authors collected clinical and other (such as genetic) data and samples, discussed the project, revised the manuscript, and provided critical feedback.

## COMPETING INTERESTS

The funders of the study had no role in the collection, analysis, or interpretation of data; in the writing of the report; or in the decision to submit the paper for publication.

SES received honoraria for serving on scientific advisory boards on biomarker testing and education for Eisai and Novo Nordisk and speaking fees from Eisai, Eli Lilly, and Novo Nordisk in 2024. SES has recently received honoraria for educational presentations from Medscape, PeerView, and the Academy for Continued Healthcare Learning. She has provided unpaid scientific advising to Acumen, Biogen, Cognito Therapeutics, Danaher, Eisai, Eli Lilly, Johnson and Johnson Innovative Medicine, Sanofi, and Siemens. GD owns stock in ANI Pharmaceuticals. JLD is an inventor on patents or patent applications assigned to Eli Lilly and Company relating to the assays, methods, reagents and / or compositions of matter for P-tau assays and Aβ targeting therapeutics. JLD has/is served/serving as a consultant or on advisory boards for Eisai, Abbvie, Genotix Biotechnologies Inc, Gates Ventures, Syndeio Biosciences, Dolby Family Ventures, Karuna Therapeutics, Alzheimer’s Disease Drug Discovery Foundation, AlzPath Inc., Cognito Therapeutics, Inc., Eli Lilly and Company, Prevail Therapeutics, Neurogen Biomarking, Spear Bio, Rush University, Washington University in St. Louis, University of Kentucky, Tymora Analytical Operations, Mindlmmune Therapeutics, Inc, Early is Good, and Quanterix. JLD has received research support from ADx Neurosciences, Fujirebio, Roche Diagnostics and Eli Lilly and Company in the past two years. JLD has received speaker fees from Eli Lilly and Company and LabCorp. JLD is a founder and advisor for Monument Biosciences and Dage Scientific LLC. JLD has stock or stock options in Eli Lilly and Company, Genotix Biotechnologies, MindImmune Therapeutics Inc., AlzPath Inc., Neurogen Biomarking, and Monument Biosciences. NCF has provided consultancy services or served on advisory boards for Abbvie, Biogen, Eisai, Eli Lilly, and Roche. AG was a consultant for Merck in 2024, as well as a member of SRB for Genentech, Muna Therapeutics and Arbor-Bio. CC is a member of the scientific advisory board of Circular Genomics and owns stocks and is on the scientific advisory board of ADmit and Alamar. CC has consulted in the last 6 months for Sanofi, NovoNordisk, and Owkin. CC has received research support from Circular Genomics, GSK, Danaher, BMS and EISAI. The rest of the authors report no conflict of interest.

## Notes

### Author Declarations

Participants were recruited through the DIAN Observational Study (ClinicalTrials.gov Identifier: NCT00869817) and have provided written consent or assent with proxy consent prior to enrollment in accordance with the latest Helsinki Declaration. The study was approved by the Human Research Protection Office and the Institutional Review Board (IRB) at Washington University in St Louis, USA and the respective participating sites.

