## SupplementaryFigures for "Abnormalities in core AD biomarkers precede inflammatory and glial markers in CSF in Autosomal Dominant Alzheimer’s Disease"

Supplementary Figures

**Supplementary Figure 1. Quality Control. A.** Principal component analysis (PCA) to identify sample outliers using NULISA quantifications. **B.** Abnormality probability curves comparing Aβ42/40 measured by IP-MS, Lumipulse and NULISA. **C.** Abnormality probability curves comparison for pTau-217 measured by IP-MS, Lumipulse, and NULISA.

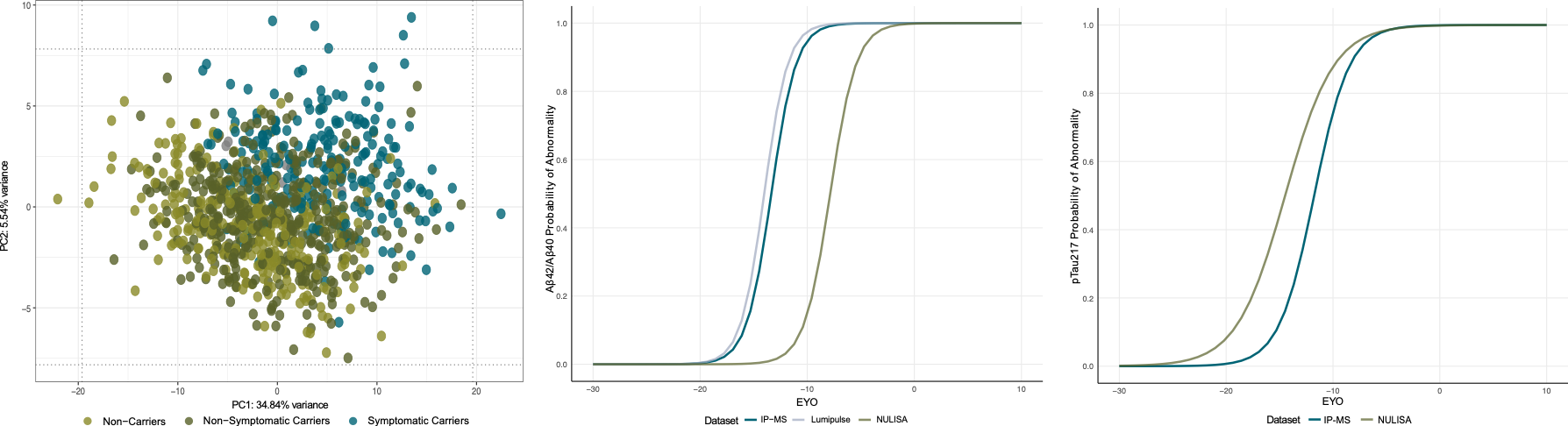

**Supplementary Figure 2** **– Distribution of CSF protein levels. A.** Density distributions of CSF pTau181 levels in mutation carriers (MC) and non-carriers (NC). The dashed line indicates the cut off. **B**. Density distributions of CSF Aβ levels in mutation carriers (MC) and non-carriers (NC). The dashed line indicates the cut-off used to define abnormal biomarker levels based on the NC distribution.

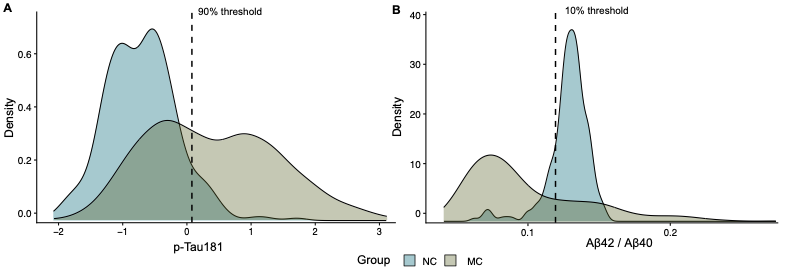

**Supplementary Figure 3** **– Differential Abundance Analysis.** **A.** Volcano plot showing the results of comparing CSF measurements in *PSEN2* mutation Carriers vs Non- Carriers (NC) at last visit. **B**. Effect size correlation for the results of the Mutation Carriers vs Non-Carriers comparison and the *PSEN2* Carriers vs Non-Carriers comparison.

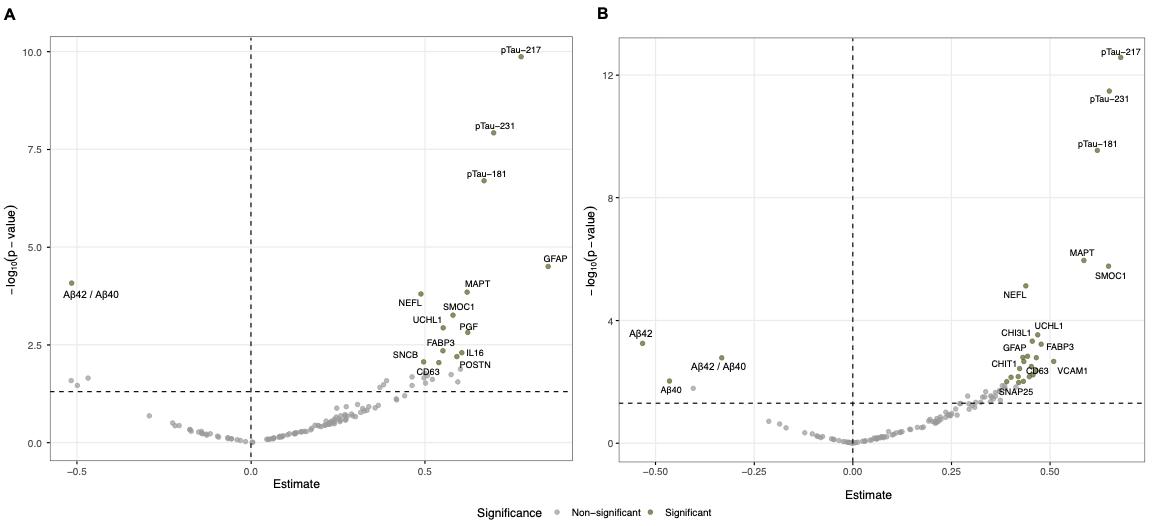

**Supplementary Figure 4** – **A.** Pairwise Spearman correlation matrix for proteins that reach at least 50% abnormality probability in mutation carriers. **B.** Gene Ontology (GO) enrichment analysis of proteins reaching abnormality, showing significantly enriched biological process.

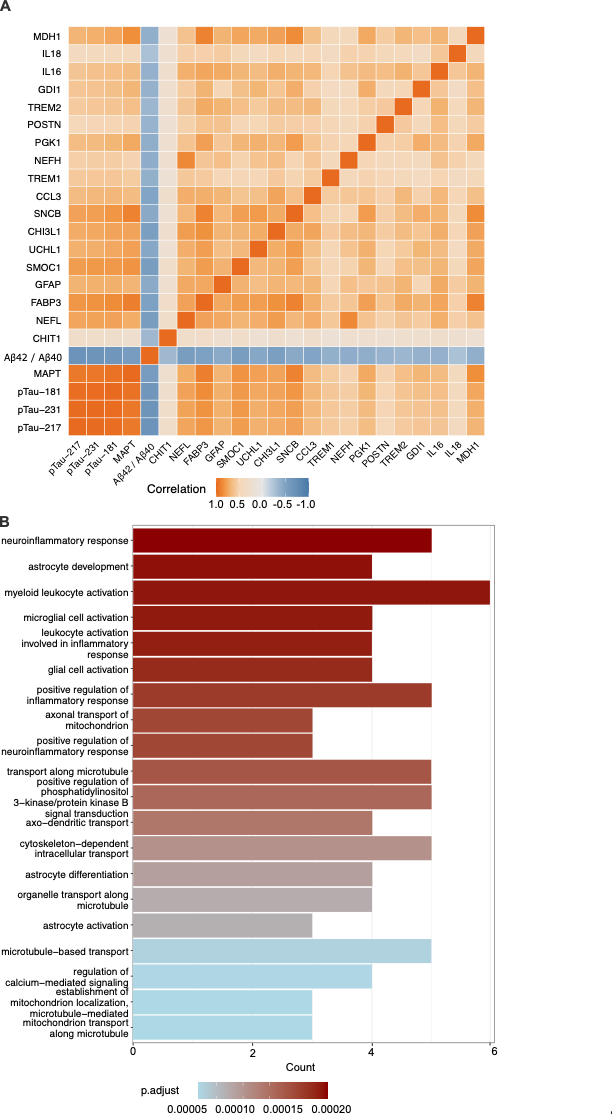

**Supplementary Figure 5** – **Multiprotein Model Design and performance for the n=24 convertors A.** Feature importance in the multi-protein model ranked by gain score. **B.** Sample distribution and performance of the single-biomarker, PET-PiB, multiprotein and age-only models across EYO windows (±2.5 years). Mean absolute error (MAE) shown with 95% confidence intervals.

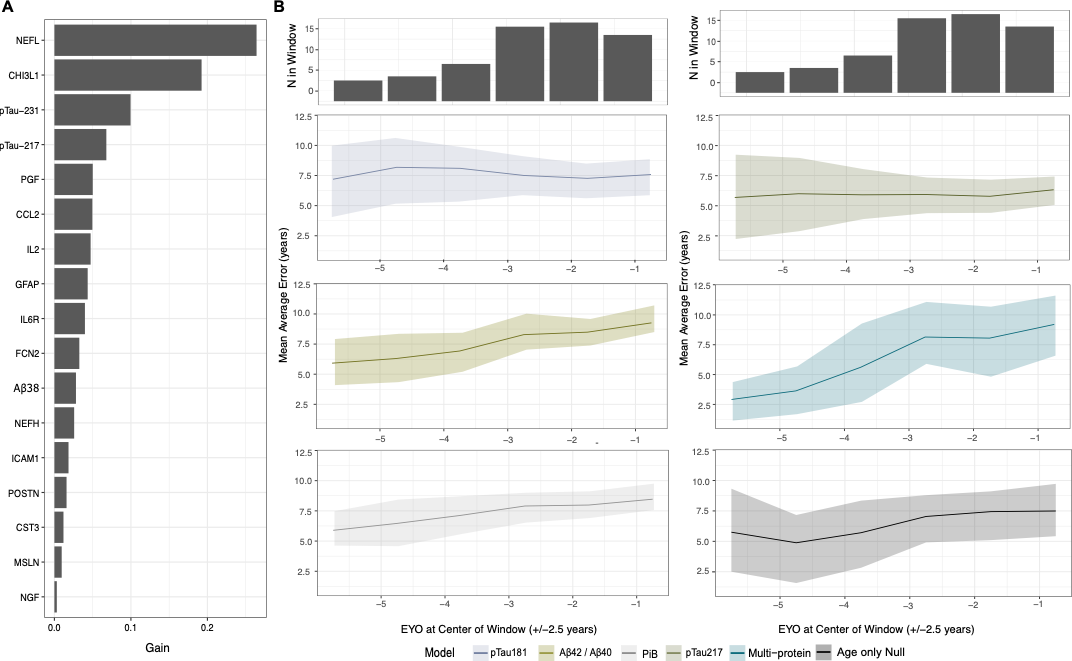

**Supplementary Figure 6** – Collapsed view of the bootstrap differences in Mean absolute error (MAE) between each model and the age-only null model in convertors (n=24) from Supplementary Figure 4. MAE shown with 95% confidence intervals.

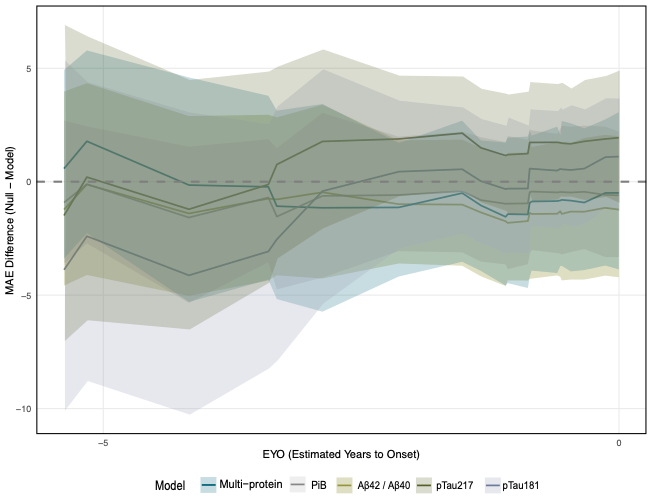
